# Anisotropy boosting improves ODF-Fingerprinting tractography in edematous brain

**DOI:** 10.1101/2025.06.18.25329353

**Authors:** Patryk Filipiak, Kamri Clarke, Timothy M. Shepherd, Mary Bruno, Dimitris G. Placantonakis, Steven H. Baete

## Abstract

Peritumoral vasogenic edema of the brain is a major confounding factor for diffusion MRI tractography. Excessive fluids accumulated in edematous white matter decrease anisotropy of water self-diffusion which affects tracking algorithms. We address this hurdle with ODF-Fingerprinting (ODF-FP) — a dictionary-based fiber reconstruction algorithm that accommodates variability of neural tissue. By adding a regularization term to the ODF-FP matching formula, we boost diffusion anisotropy to improve white matter fiber identification in edematous regions.

## Purpose

Peritumoral vasogenic edema of the brain is a major confounding factor for diffusion MRI (dMRI) tractography^[1]^. Excessive fluids accumulated in the extra-axonal space decrease diffusion anisotropy, causing premature termination of reconstructed White Matter (WM) fibers^[2]^. As a result, tractography images are often inaccurate in proximity to tumor mass, which hampers resection planning.

We address this hurdle with ODF-Fingerprinting (ODF-FP)^[3]^ — a dictionary-based fiber reconstruction algorithm that accommodates variability of neural tissue^[4]^. By adding a regularization term to the ODF-FP matching formula, we boost diffusion anisotropy to improve WM fiber tracking in edematous brain.

## Theory

Let *𝒟* be an ODF-dictionary. We define the ODF-FP matching formula:

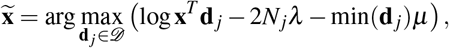

where **x** is an input ODF, **d** _*j*_ ∈ *𝒟* are dictionary ODFs (each with *N*_*j*_ ≥ 0 crossing fibers) among which the best matching candidate 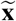 is selected, and *λ*,*μ* ≥ 0 are regularization weights. Note that the added term min(**d** _*j*_) boosts diffusion anisotropy by penalizing the lowest values of ODF and is intended to use only in edema regions (i.e.,*μ* = 0 in the remaining voxels).

## Methods

We considered dMRI of 10 brain tumor patients (51± 11 y/o) with vasogenic edema infiltrating the Arcuate Fasciculus (AF; 9 cases) or the Corticospinal Tract (CST; 7 cases). The images were acquired with a 3T Siemens Prisma (Erlangen, Germany) MR scanner at 2 × 2 × 2 mm^3^, *TE/TR* = 92*/*5900 ms, 60 directions at *b* = 300, 1100, 2500, 5000 s/mm^2^ and 8 at *b* = 0. Our postprocessing in MRtrix3^[5]^ included denoising, Gibbs ringing removal, correction of B1 field inhomogeneity and eddy currents. We then executed ODF-FP with a dictionary of 10^6^ items having 0 ≤*N*_*j*_ ≤3 crossing fibers per voxel, *λ* = 10^−5^, and an exploratory set of anisotropy boost factors*μ* ∈ { 0, 0.01, 0.02,…, 0.30} applied in manually drawn edema regions. For comparison, we processed the same images using: Constrained Spherical Deconvolution with Multi-Shell Multi-Tissue option (CSD MSMT)^[6]^, Free Water Elimination Diffusion Tensor Imaging (FWE DTI)^[7]^, Freewater estimatoR using iNtErpolated iniTialization (FERNET)^[8]^, Functional magnetic resonance imaging of the brain Software Library (FSL) Bedpostx^[9]^, and Generalized Q-sampling Imaging (GQI)^[10]^. For each method *m*, we dissected AF and CST using automated tracking in DSI Studio^[11]^, then calculated the respective overlap enhancements (OE) defined as normalized volumes (vol):

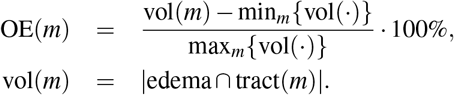

## Results

Linear increment of the anisotropy boosting factor*μ* gradually increased OE of ODF-FP, then reached plateau within the 0.10–0.20 range (Figure 1). The improvement in shape of the reconstructed tracts was particularly visible in AF (Figure 1A). Quantitatively, ODF-FP with the proposed regularization term (weighted by*μ* = 0.15) considerably outperformed other tested methods (Figure 2).

**Figure 1:**
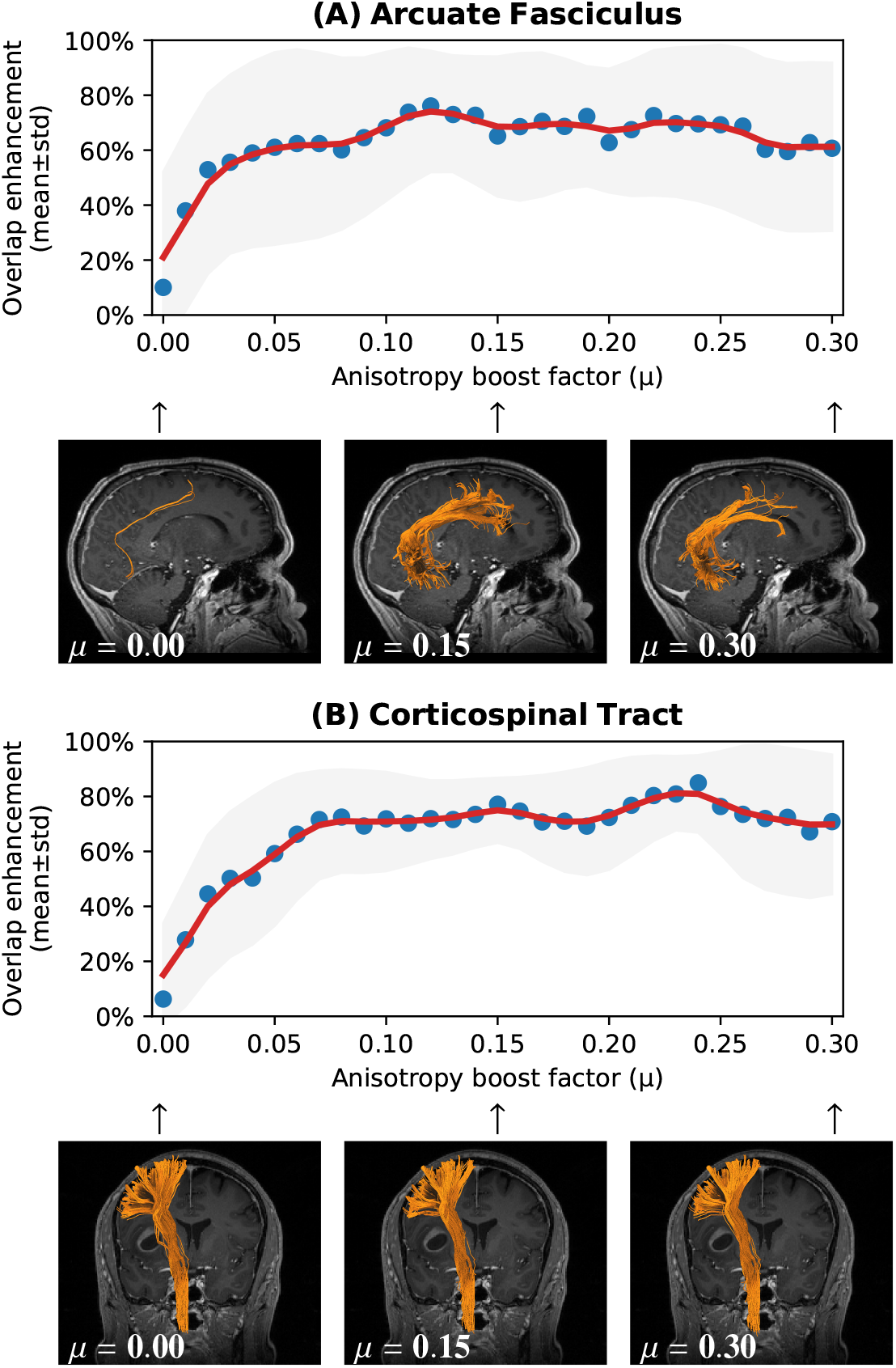
Mean overlap enhancements in ODF-FP reconstruction of (A) Arcuate Fasciculus and (B) Corticospinal Tract with a range of anisotropy boost factors*μ*.

**Figure 2:**
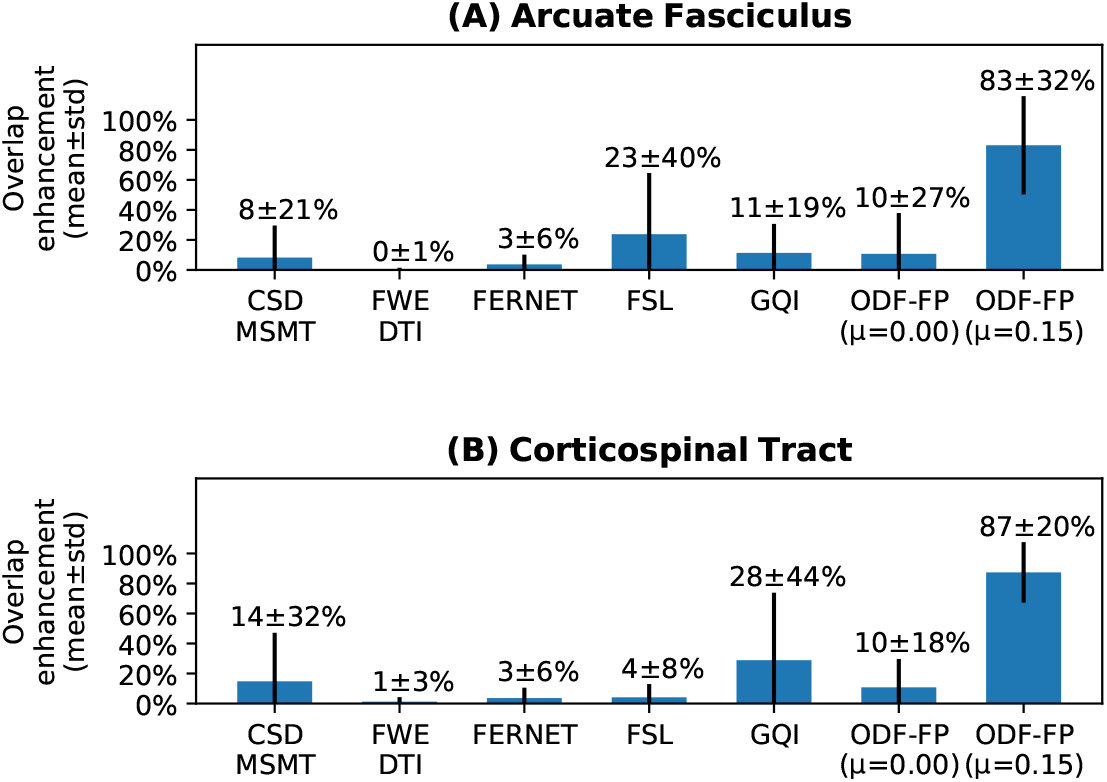
Comparison of mean overlap ehancements in all tested methods.

## Discussion and Conclusion

ODF-FP with the proposed modification to boost diffusion anisotropy has the potential to overcome dMRI signal distortion due to vasogenic edema. Future work should address automatic recognition of edema regions and adaptability of ODF-FP to clinically feasible dMRI acquisition protocols.

## Data Availability

All data produced in the present study are available upon reasonable request to the authors.

## Acknowledgements

This project was supported in part by the National Institutes of Health (NIH: R01-EB028774, R01-NS082436) and performed under the rubric of the Center for Advanced Imaging Innovation and Research (CAI^2^R, https://www.cai2r.net), an NIBIB Biomedical Technology Resource Center (NIH P41-EB017183).

